# Palmar cooling mitigates exercise-induced immune suppression after high-intensity training: A randomized trial

**DOI:** 10.1101/2025.11.05.25339605

**Authors:** Amy S. Tsai, Vinh Cao, Romain Lagarde, Sasha Anronikov, Colin Liu, Kotaro Miyazaki, Maximilian Sabayev, Sam Moghaddam, Dorien Feyaerts, Jakob Einhaus, Dyani Gaudilliere, Ed Ganio, Charles Darracott, Maigane Diop, Amelie Cambriel, H Craig Heller, Brice Gaudilliere

**Affiliations:** Stanford School of Medicine, Department of Surgery, Division of General Surgery, Stanford, CA; Stanford School of Medicine, Department of Anesthesiology, Stanford, CA; Stanford University, Department of Biology, Stanford, CA; Stanford School of Medicine, Department of Surgery, Division of Plastic and Reconstructive Surgery, Stanford, CA; Institute for Computational and Mathematical Engineering, Stanford University, USA

## Abstract

**Objectives:** We investigated whether palmar cooling alters inflammatory responses following a single session of high-intensity eccentric exercise. We hypothesized that palmar cooling during rest intervals would attenuate maladaptive inflammatory responses while preserving beneficial immune adaptations necessary for muscle repair and recovery.

**Methods:** In this randomized interventional study, 20 healthy adults were matched by sex and one-repetition maximum (1RM) for bicep curls. Participants performed 10 sets at 70% 1RM, receiving either palmar cooling at 14ºC or a thermoneutral control at 30ºC during 3-minute inter-set rest periods. Whole blood was collected at baseline, immediately post-exercise, and on post-exercise days 1, 2, and 4 for comprehensive immune profiling with mass cytometry. Blood lactate and pain scores were also recorded.

**Results:** Exercise induced broad immunosuppression that was significantly attenuated in the palmar cooling group by 2 days post-exercise (AUC=0.79, *p*=0.03). The cooled group had decreased immunosuppressive activity and increased inflammatory innate immune mechanisms in the cooled group. Palmar cooling also significantly reduced lactate levels compared to controls (*p*=0.024).

**Conclusion:** Palmar cooling at 14ºC during rest intervals effectively modulated the immune response to high-intensity exercise and reduced lactate accumulation. These findings suggest palmar cooling may serve as a promising intervention to mitigate exercise-induced immunosuppression and support recovery.

**Trial identifier:** NCT07215338 (retrospectively registered).

**Summary Box:** *What is already known on this topic:* *High intensity eccentric exercise contributes to physiological heat stress which induces significant immune alterations that can lead to a transient period of immunosuppression and increased incidence of respiratory tract infections*.

*What this study adds:* *Palmar cooling resulted in cell-type-specific alterations in innate and adaptive immune cell signaling activity, particularly decreasing immunosuppressive cell responses while enhancing inflammatory innate immune mechanisms implicated in pathogen defense*.

*How this study might affect research, practice or policy:* *Palmar cooling may serve as an effective intervention for managing exercise-induced immunosuppression and supporting recovery in athletes. Integration of palmar cooling techniques into athletic training regimens and rehabilitation protocols can ultimately enhance exercise performance and reduce the adverse effects associated with high-intensity exercise*.

## INTRODUCTION

High intensity exercise imposes substantial physiological stress that mobilizes the immune system, leading to a transient leukocytosis immediately post-exercise followed by leukopenia prior to returning to baseline[1]. This transient immunosuppression, the “open window,” spans approximately 3–72 hours post-exercise, during which host pathogen defense is impaired[2]. Although endurance athletes have higher rates of respiratory illness that underscore this vulnerability[3–8], post-exercise immune alterations also serve beneficial purposes, playing an essential role in muscle repair and recovery after exercise[9,10]. These same inflammatory pathways underlie the physiology of delayed-onset muscle soreness (DOMS)[11,12]; when this inflammatory response becomes excessive or prolonged, athletes face impaired recovery, ultimately limiting capacity for training and performance. These competing responses highlight the need for strategies that can temper dysregulated inflammation while preserving mechanisms involved in tissue healing and exercise endurance.

The impact of immunological stress on recovery is further complicated by thermoregulatory strain. Heat stress is a major contributor to impaired performance and range from mild (heat cramps or exercise associated muscle cramps) to severe (heat exhaustion and heat stroke)[13] and can severely damage the innate immune system as well as increase anti-inflammatory cytokine production[14]. Given the intertwined nature of thermal and immune responses, strategies that modulate body temperature during exercise may also influence post-exercise inflammation. We hypothesize that palmar cooling, a non-pharmacologic technique used to regulate body temperature during exercise, can mitigate the inflammatory responses without compromising the beneficial adaptations of exercise. Previous studies have explored the effects of palmar cooling on athletic performance with findings of increased work volume[15], reduced fatigue in high-intensity bench press exercises[16], and improved aerobic endurance[17]. These effects are additive and can be sustained for up to six weeks after intervention[15] suggestive of systemic modulation of human physiology. However, the effects of palmar cooling on the systemic immune response to high intensity exercise remain largely unexplored.

To address this gap, we employed mass cytometry, a high-dimensional single-cell analysis technology that quantifies over 40 protein markers per cell and has been used to investigate the complex dynamics of the immune system in response to physiological stress[18–26]. In this study, we leveraged mass cytometry to perform a comprehensive, time-dependent and functional analysis of the human immune response elicited by intense exercise and to determine how palmar cooling modulates these responses.

## METHODS and MATERIALS

### Study Design

#### Participants

This prospective randomized clinical study was approved by the Institutional Review Board (IRB) of Stanford University School of Medicine. Participants were recruited by flyers and were introduced to the study through a written consent form approved by Stanford’s IRB. Inclusion criteria included 18 years of age or older and absence of underlying immune disorders. Exclusion criteria included inability to lift greater than 35 pounds, or the weight of the cross bar. 40 individuals responded to recruitment flyers, 22 consented, and 20 enrolled and completed the study, 20 were included in the primary mass cytometry analysis and 22 included in secondary outcome analysis. Two were excluded from mass cytometry analysis given inability to obtain blood samples for the baseline or any timepoints on the day of the exercise trial. CONSORT map is depicted in Supplemental Figure 1. The trial was registered retrospectively, and no changes to the protocol were made after enrollment began.

#### Patient and Public Involvement

Patient and public involvement (PPI) was integrated from the onset of the study to ensure the research was aligned with participant priorities and experiences. Feedback from student athletes was crucial in designing the study protocol, particularly in determining the feasibility and accessibility of the palmar cooling intervention. Recruitment for the study was conducted at local gyms and dorms within Stanford University, where patients and the public were first involved during the development of the research questions and outcome measures. During recruitment, participants discussed and agreed to the burden of the intervention (one-hour exercise session) and the time required for participation (five phlebotomy sessions, one prior to exercise and four subsequently, with self-recorded logs of pain scores), ensuring the study was respectful of their commitments. Participants agreed to share de-identified demographics and biological data with the public.

#### Equity, Diversity and Inclusion in Research

To address diversity in our study population, recruitment efforts were strategically planned to ensure a balanced representation. Recruitment was conducted at various locations within and outside of Stanford University, including gyms and dorms, to reach a diverse cross-section of the student body and local community. Efforts were made to guarantee sex balance among participants, and outreach initiatives at gyms outside of Stanford University were designed to engage individuals from different socioeconomic and education backgrounds. Sex-equality was controlled for in all subsequent analysis for interpretability and generalizability.

Diversity was ensured among the investigator and author team by including members from different sexes, academic levels, and disciplines. The multidisciplinary team was comprised of researchers (undergraduate students, graduate students, postdoctoral fellows, senior scientists and expert professors) with diverse backgrounds in clinical medicine, biostatistics, physiology, immunology and athletics.

#### Bicep curls

Participants were paired by sex, age and two-arm bicep curl 1 repetition maximums (1RM). Control participants performed 10 sets bicep curls at 70% of their 1RM until muscle failure. Cooled participants performed the same number of repetitions in each set as their control counterparts.

#### Randomization and Blinding

Once individuals were matched, group allocations were determined with a coin toss.

#### Palmar cooling

Subjects randomized into the palmar cooling arm received three minutes of cooling at a heat sink temperature of 14ºC between sets. Control participants received three minutes of the same treatment at a heat sink temperature of 30ºC. Each subjects placed their palms on 2 cylindrical 8 by 6.5 inches (flatten) poly-urethane water perfusion pads for palmar cooling. The perfusion pads were plumbed to a Thermo Scientific Haake SC150 water bath with a flow rate of 4 L/min. Two water baths were used in the study, one set to 14ºC and the other to 30ºC

#### Lactate measurement

Blood lactate was measured using Lactate Plus test strips and meter from Nova Biomedical. Blood lactate was measured pre-exercise and post-set-10 of bicep arm curl.

### Outcomes

#### Primary outcome

The primary outcome is specified by modulation of immune cell proinflammatory signaling responses in the intervention arm.

#### Secondary outcomes

Secondary outcomes included differences in lactate levels prior to and immediately following exercise, and pain scores according to the Borg Perceived Pain Scale [27] for five days following exercise. Delayed onset of muscle soreness (DOMS) was defined as a pain score >5 on PED2.

### Sample processing for mass cytometry analysis

#### Whole blood processing

Whole blood samples were collected in sodium-heparin tubes prior to exercise (Baseline, BL), on day of exercise (DOE) immediately following exercise, and post-exercise days (PED) one, two, and four. All samples were processed within 30 minutes of collection, stimulated with 1ug/mL of lipopolysaccharide (LPS), 100ng/mL of Interferon-α (IFNα) or left unstimulated to measure endogenous activity, and then fixed with Smart Tube buffer (Smart Tube Inc, San Carlos, CA) and stored at -80ºC until ready for mass cytometry analysis.

#### Sample processing for mass cytometry analysis

Samples were thawed, and erythrocytes lysed using Thaw-Lyse Buffer (Smart Tube Inc., San Carlos, CA), prior to barcoding with palladium isotopes, and staining with thirty-seven metal-conjugated surface and intracellular antibodies in accordance with prior established protocols ([22,24], Supplemental Table 1). In summary, nine palladium and platinum isotopes (^104^Pd, ^105^Pd, ^106^Pd, ^108^Pd, ^110^Pd, ^194^Pt, ^195^Pt, ^196^Pt and ^197^Pt) were used in combinations of four to barcode and run 96 samples per mass cytometry plate. To maximize the sensitivity of the assay to detect differences between treatment and control samples, paired longitudinal samples were run simultaneously using the Helios mass cytometry system (Standard BioTools, South San Francisco, CA). FCS files were normalized [28] and de-barcoded 11/5/2025 9:40:00 AM.

#### Manual gating

Manual gating was performed using CellEngine (CellEngine, Fremont, CA). The gating strategy can be found in Supplemental Figure 2). Seventeen cell-types were included in all subsequent analysis: neutrophils, CXCR4^+^ neutrophils, B-cells, CD56^+^CD16^-^ Natural Killer (NK) cells, CD56^lo^CD16^+^ NK cells, CD4^+^CD45RA^-^ T cells (CD4^+^ T_mem_), CD4^+^CD45RA^+^ T cells (CD4^+^ T_naive_), CD25^+^FoxP3^+^CD4^+^ T cells (T_regs_), CD8^+^CD45RA^-^ T cells (CD8^+^ T_mem_), CD8^+^CD45RA^+^ T cells (CD8^+^ T_naive_), TCRγδ T cells, CD14^+^CD16^-^ classical monocytes (cMCs), CD14^-^CD16^+^ non-classical monocytes (ncMCs), CD14^+^CD16^+^ intermediate monocytes (intMCs), monocytic myeloid-derived suppressor cells (M-MDSCs), myeloid dendritic cells (mDCs), and plasmacytoid dendritic cells (pDCs).

### Statistical analysis

*Power Analysis*. Based on previous data documenting the activation of STAT3 and MAPK signaling pathways in MDSCs 24 hours after surgery [18] we estimated that a sample size of 8 participants in each group would be sufficient to provide 95% power at P < 0.05 to detect a >40% change in STAT3 phosphorylation in monocyte subsets (Cohen’s d effect size of 1.96) following exercise using student’s t-statistic. This power analysis was performed using G*Power [29].

#### Cell frequency

Cell frequency was calculated as a percentage of live mononuclear (CD45^+^CD66^-^cPARP^-^) for mononuclear cell subsets and as a percentage of live leukocytes (cPARP^-^) for neutrophils at each time point. Cell frequencies were calculated at each time point.

#### Functional markers

Functional markers were expressed as an arcsinh transformed value of the median for unstimulated samples while stimulated samples (LPS, IFNα) were expressed as an arcsinh ratio from the unstimulated sample. The following functional markers were included: phospho-(p)STAT1, pSTAT3, pSTAT5, pSTAT6, pNFκB, pMAPKAPK2, pP38, prpS6, pERK1/2, pCREB, and IκBα. Functional markers expressions were calculated at each time point.

#### Visualization of immune cell atlas and Bootstrapped clustering methods

A correlation matrix was generated using Spearman’s method, and a network graph was constructed where nodes represented features and edges represented strong correlations (absolute value > 0.8). The node attributes included color (based on normalized differences) and size (based on -log10 of p-value). The network was visualized using a Kamada-Kawai layout, with significant nodes labeled.

#### Univariate analysis

Univariate p-values for individual immune features (frequencies and functional responses) and clinical data (lactate levels and pain scores) were obtained using a Wilcoxon test. The differences between timepoints were calculated using Analysis of Variance (ANOVA). There were no missing data in this dataset. All statistical analysis was reviewed to be consistent with the CHAMP guidelines[30].

#### Multivariable Analysis

A knowledge-based penalization was applied for each stimulation (Unstim, LPS, IFNα, Supplemental Table 2). Multivariable sparse models were then used to classify samples at a given timepoint between the case and control groups. Multiple models were considered: Least Absolute Shrinkage and Selection Operator (LASSO)[31], Adaptive LASSO[32], as well as their Stabl[33] counterparts. Each model was built on all the data as once (Early Fusion), or on each stimulation separately, with the predictions then unified in a final model (Late Fusion). Model performance was estimated via Leave-One-Out cross-validation. For each timepoint classification task, the best model was determined by best Area Under the Receiver Operating Characteristic (AUROC). The best model’s features were then reported (Supplemental Tables S4, S5, S6). In the same manner, models were fit to classify samples between baseline and a given timepoint.

## RESULTS

### Clinical characteristics of participants

Twenty sex-matched participants were paired on two-arm biceps curl one-repetition maximums (1RM). Control participants performed 10 repetitions of bicep curls at 70% of their 1RM until muscle failure. Cooled participants performed the same number of repetitions as their control counterparts. During the three-minute rest periods between sets, all participants placed their hands in a palmar cooling interface. For control participants, the temperature was set to 30ºC while for cooled participants, the temperature was set to 14ºC. Whole blood was obtained at five timepoints including baseline prior to intervention (BL), immediately after exercise (Day of Exercise, DOE), and on Post-Exercise Days (PED) one, two and four. Samples were either left unstimulated to measure endogenous activity or stimulated with exogenous inflammatory ligands including Lipopolysaccharide (LPS, a bacterial endotoxin that drives severe infection and fever responses), and Interferon-α (IFNα, which mediates the body’s response to viral infections and malignancy) and measured using mass cytometry. Multivariable analysis performed using Stabl[33]. Point of care lactate levels were obtained prior to exercise on day of exercise and immediately after last set of bicep curls and palmar intervention. Lactate levels in the control arm increased significantly more than the cooled arm (1.2±1.4 [95% CI], *p*=0.024). This difference was more pronounced in female (1.4±1.2, *p*=0.018) compared to male participants (1.1±3.0, *p*=0.50) (Figure 1C, Supplemental Figure S3). Participants were asked to record their pain scores daily for five days after exercise. Delayed onset of muscle soreness (DOMS) was defined as pain > 5 on PED2. No individuals met the definition of DOMS in this study. Pain scores did not differ between the participants (Supplemental Figure 4). Study design is depicted in Figure 1.

**Figure 1.**
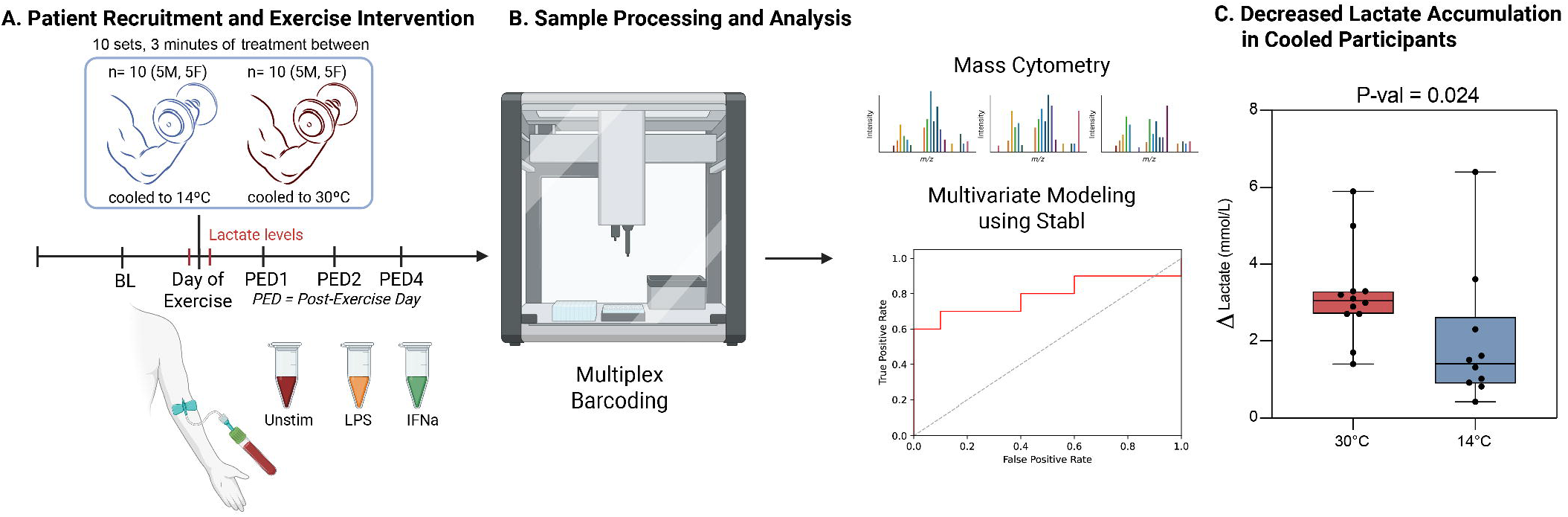
Study Design. (A) 20 individuals were matched by sex and two-arm bicep curl one-repetition maximums (1RM). Whole blood samples were collected prior to exercise, immediately following exercise, and on Post-Exercise Days (PED) 1, 2, and 4. Samples were stimulated with exogenous LPS, IFNα, or left alone. (B) Samples were analyzed simultaneously using multiplex barcoding and mass cytometry. Multivariable modeling was performed using Stabl. (C) Significant differences in lactate levels after exercise intervention (*p-val* = 0.024) Change in lactate levels are depicted in mmol/L. P-values are calculated using two-sided Wilcoxon rank-sum test. All boxplots show median values, interquartile range, whiskers of 1.5 times interquartile range. Created with the aid of BioRender.

### Assessment of the human immune response to exercise

Exercise induced substantial alterations of the immune system that spanned multiple innate and adaptive immune cell subsets. Immune responses induced by exercise were visualized on a correlation network highlighting cell-type- and signaling-specific differences at each post-exercise time point relative to baseline (Figure 2). Multivariable modeling of the mass cytometry data using sparse machine learning (Stabl, [33]) identified immune-system wide differences on PED1 (AUC = 0.80, *p*=0.0015), PED2 (AUC = 0.69, *p*=0.041) and PED4 (AUC = 0.69, *p*=0.040) but not on DOE (AUC=0.67, *p*=0.064, Supplemental Table 3), compared to baseline.

**Figure 2.**
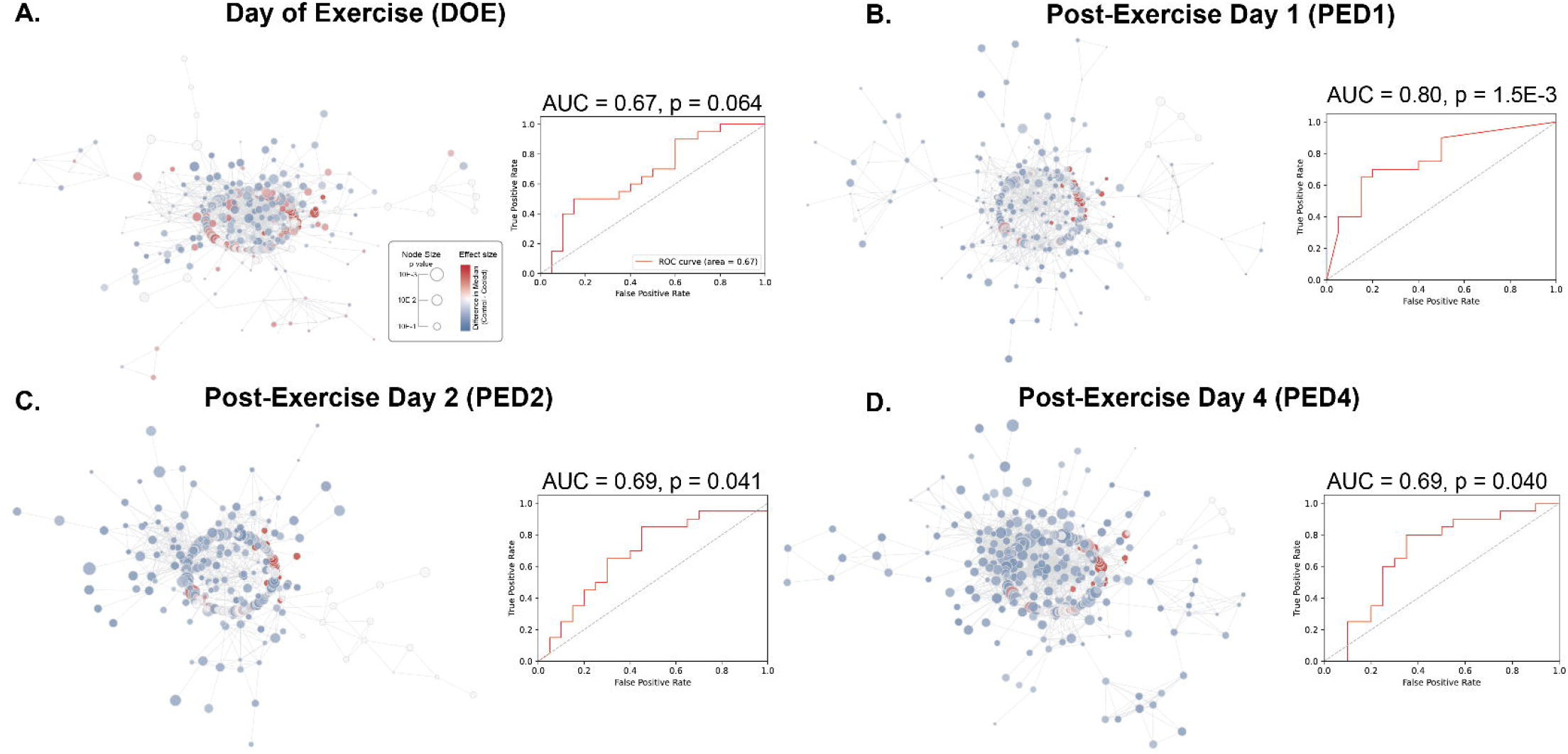
Single-cell analysis of the peripheral immune response to exercise with mass cytometry. Correlation networks (left panels) and multivariable modeling (right panels) depicting the peripheral immune response to exercise on (A) Day of Exercise (DOE), (B), Post-Exercise Day (PED) 1, (C) PED 2, and (D) PED Day 4. Left panels: Correlation networks. Each node represents one immune feature (population, functional marker, stimulation). Node color represents directionality of change (Blue = decreased in response to exercise, Red = increased). Node size corresponds to -log(*p-val*) of each feature. Right panels: multivariable modeling (Stabl) differentiates immune response on DOE (AUC = 0.67, *p-val* = 0.064), PED1 (AUC = 0.80, *p-val* = 0.0015), PED2 (AUC = 0.69, *p-val* = 0.041) and PED4 (AUC = 0.69, *p-val* = 0.040) from baseline. Model performances were evaluated using cross-validation.

To gain mechanistic insight into the multivariable analysis, we examined immune features that were shared across the time-dependent models. We observed a rapid increase in the frequency of neutrophils and monocyte subsets immediately following exercise that was sustained at all time points (Fig 3, Supplemental Figure 5, Table 4). Interestingly, monocyte subsets exhibited diminished responses to inflammatory stimulations, including the pERK, pCREB and pP38 response to LPS, and the pP38 response to IFNα. We also observed a reduced pP38 response to IFNα in non-classical monocytes and B-cells, suggestive of exercise-induced immunosuppression.

**Figure 3.**
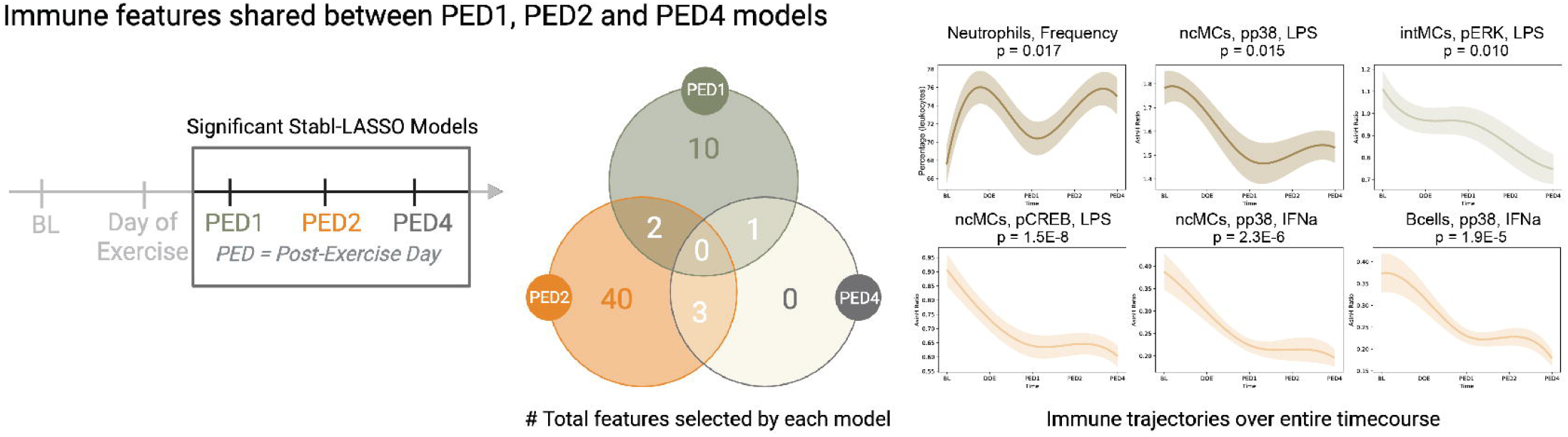
Immune features shared between PED1, PED2 and PED4 models. (Left) Timeline of samples collected, significant (*p-val* < 0.05) models are boxed. (Center) Total number of features selected by each model depicted in a venn diagram. (Right) Individual trajectories of features that were selected by at least 2 models are depicted and colored according to venn diagram. Neutrophil frequency depicted as percentage of total leukocytes and intracellular functional markers are depicted as median AsinH ratio value with standard error. ANOVA p-values are provided for each feature.

### Modulation of immune trajectories in response to palmar cooling

Having defined the effects of high-intensity exercise on peripheral immune cells, we tested whether palmar cooling modulates this immune response. We performed a multivariable analysis comparing the mass cytometry data at each time point between subjects receiving 30ºC or 14ºC palmar cooling. Significant differences in the immune response to exercise were observed between the groups on PED2 (AUC 0.79, *p*=0.03, Fig. 4a). No differences between the two groups were identified at baseline, prior to exercise and palmar cooling (AUC 0.63, *p*=0.34), immediately after exercise (AUC 0.58, *p*=0.57, Day of Exercise), on PED1 (AUC 0.59, *p*=0.52), or on PED4 (AUC 0.71, *p*=0.12, Supplemental Table 4). These results indicate that palmar cooling transiently modulates the cellular immune response to exercise.

**Figure 4.**
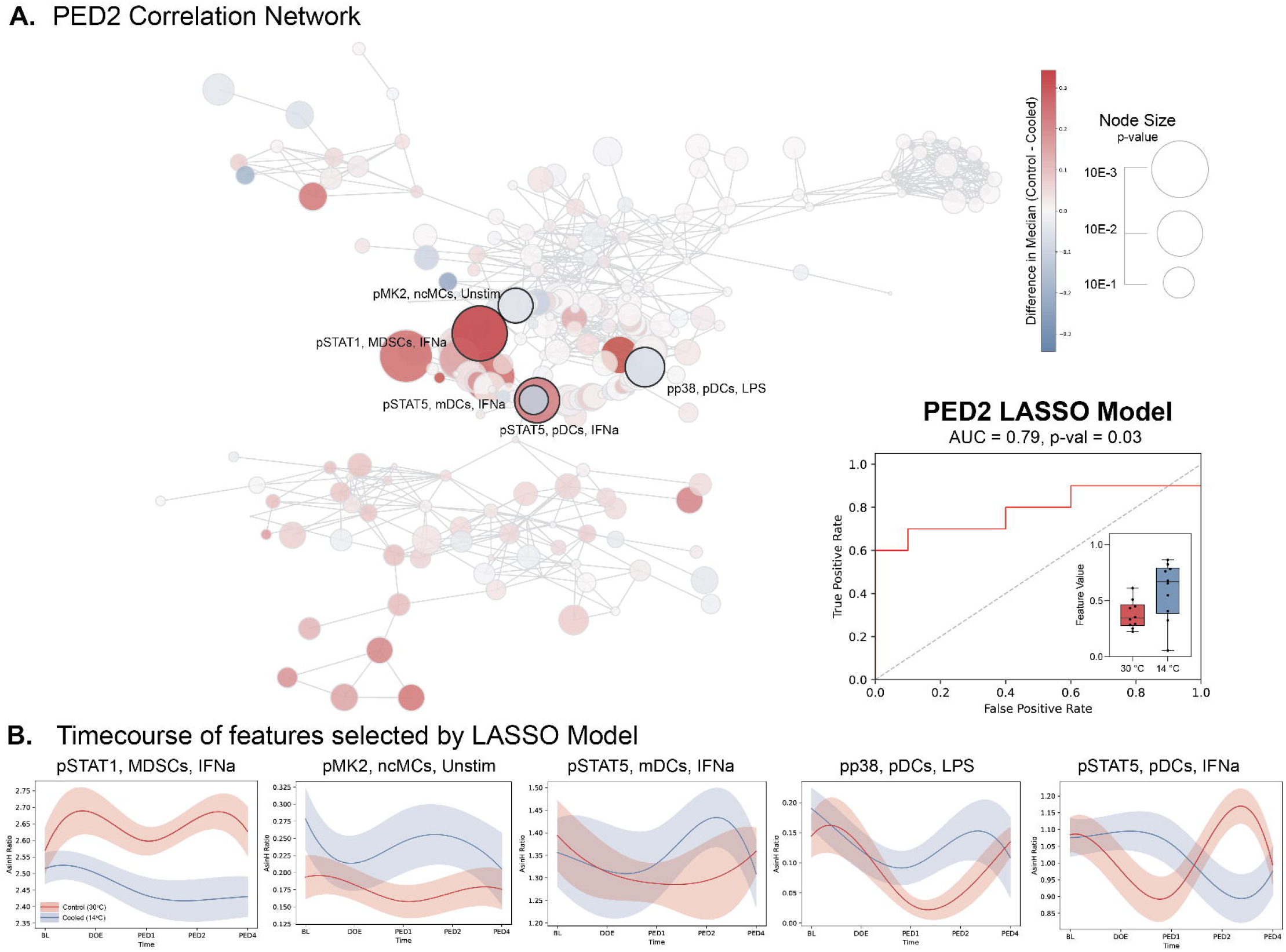
Immune modulations in response to palmar cooling. (A) Correlation network (left) depicting immune difference between Control and Cooled arms on Post-Exercise Day 2. Each node represents one immune feature (population, functional marker, stimulation). Node size corresponds to -log(*p-val*) of each feature. Node color represents directionality of change in cooled participants compared to control (Red = higher in Control, Blue = higher in Cooled). Features selected by the model are labeled. The Stabl-LASSO model (right) achieved an AUC of 0.79 (*p-val*=0.03). Model performances were evaluated using cross-validation. (B) Weighted features selected by the LASSO model are depicted. Divergent trajectories of features in each arm (Red = Control, Blue = Cooled) are depicted with median and SE across time.

Examination of the most robust model features that differentiated the cooled from the control groups at PED2 over time provided a biological interpretation of the multivariable analysis (Fig. 4b). We observed that pSTAT1 activity quickly diverged after exercise and was lower in the immunosuppressive MDSCs in response to IFNα in the cooled arm, while inflammatory pathway markers such as pMK2, pp38 and, pSTAT5 were higher in response to stimulation in mDCs and monocytic cell populations. These responses started on PED1 and became more pronounced by PED2 before largely returning to baseline by PED4. Conversely, pSTAT5 activity in pDCs was initially elevated after exercise on PED1 but then became significantly lower in the cooled arm in response to IFNα stimulation.

## DISCUSSION

In this study, we employed high-dimensional immune monitoring with mass cytometry to comprehensively characterize the human immune response to high-intensity exercise. Exercise induced profound, cell-specific immune responses that emerged by 24 hours post-exercise and persisted for up to four days. Furthermore, we found that palmar cooling modulated this immune response, decreasing immunosuppressive MDSC activity and increasing pSTAT5 and pMK/p38 phosphorylation in pro-inflammatory innate immune cells, thereby restoring immune balance. These findings provide mechanistic insight into the immunological effects of exercise and identify palmar cooling as a potential strategy to mitigate maladaptive post-exercise immunosuppression in athletes and improve recovery following intense exercise[3–8].

High dimensional analysis of the mass cytometry dataset showed that exercise orchestrates a broad yet coordinated immune recalibration. This immune response was particularly pronounced within innate immune compartments. Specifically, exercise resulted in reduced pCREB, pERK, and pP38 signaling activity in neutrophils and monocytes following stimulation with exogenous LPS. These results corroborate previous findings of a transient post-exercise immunosuppressive “open-window” [1,2], which is more pronounced in innate than adaptive immune cell subsets[34]. Our findings also align with prior reports of post-exercise leukocytosis followed by functional hyporesponsiveness to exogenous bacterial LPS stimulation, consistent with previously described extravasation of marginated neutrophils with diminished effector response[35]. Interestingly, we also observed alterations of adaptive immune cell function, including a blunted B cell response to exogenous IFNα, which has not been previously described. Given the critical role of B cell-mediated humoral immune response in immunity against pathogens, decreased B-cell responses could contribute to the increased susceptibility of performance athletes to infection in the days following intense exercise, such as upper respiratory tract infections[6–8]. Accordingly, the immune responses elicited by high-intensity exercise resemble those triggered by acute injury, engaging both innate and adaptive pathways that orchestrate key recovery processes such as pathogen defense and wound healing[19,21,22,25,26,36].

Comparison of volunteers exposed to 14°C palmar cooling with controls revealed that cooling modulated exercise-induced immune responses, with the most pronounced differences observed on PED2. The timing of this effect aligns with the typical onset of exercise-induced immunosuppression and DOMS. Cooling was associated with a decrease in the activity of immunosuppressive cells, such as MDSCs, and an increase in the activity of pro-inflammatory innate immune cells, including DCs and monocytes, restoring responsiveness toward pre-exercise baselines. Interestingly, we observed a divergent response of pDCs to bacterial (LPS) versus viral (IFNα) perturbation. pDCs, a rare but critical component of the human innate immune response, recovered more rapidly to LPS stimulation in cooled participants compared with controls, but exhibited a diminished response to IFNα. Since pDCs express low levels of TLR4, they can respond directly to LPS in a manner resembling pro-inflammatory classical DC responses. By contrast, circulating IFNα blocks the ability of pDCs to undergo this inflammatory “fate-switch” [37], instead promoting a tolerogenic (anti-inflammatory) phenotype, a phenomenon also reported in the context of chronic exercise [38,39]. Palmar cooling may enhance pro-inflammatory responses to LPS while attenuating anti-inflammatory responses to IFNα, potentially contributing to restoration of pre-exercise immune baseline. Together, these findings support the hypothesis that palmar cooling can reverse exercise-induced immunosuppression while preserving beneficial inflammatory dynamics.

Palmar cooling also significantly reduced lactate accumulation, consistent with previous studies[40–42]. Lactate plays a dual role in immunity: transient increases trigger beneficial repair pathways, whereas sustained elevations, as seen in chronic disease, drive persistent suppression[43]. Lactate has been correlated to the degree of exercise exertion and aerobic capacity, as measured by oxygen consumption[40]. By lowering post-exercise lactate accumulation, palmar cooling may improve both endurance performance [15,17] and immune homeostasis. Given lactate’s role in modulating pathways such as NFκB and MAPK[44], these findings provide a unifying mechanism for dual metabolic and immunologic benefits of palmar cooling.

### Clinical Implications

These findings have direct relevance for athletic performance and health. High-performing endurance and high-load athletes are particularly vulnerable to post-exercise immunosuppression and infections (such as respiratory illness), which can disrupt training schedules and impair recovery. By attenuating maladaptive immunosuppression and reducing lactate accumulation, palmar cooling may offer a practical, non-pharmacologic intervention to maintain immune competence in the critical post-exercise “open window” period. This approach could be integrated into training and recovery protocols for athletes, especially during periods of high training load, competition, or travel where infection risk is heightened. Beyond athletics, the ability of palmar cooling to simultaneously modulate immunity and metabolism suggests broader translational applications—for example, in rehabilitation, occupational settings involving extreme exertion, or clinical populations where exercise-induced immune dysfunction may pose added risks.

### Limitations

This study is subject to several inherent limitations. Given the nature of the investigation, participants were unable to be entirely blinded as to which treatment arm they had been randomized to, which may have led to observer bias. Additionally, the controlled model of muscle stress simulated by this single exercise session with cooling may not capture the full complexity of endurance or multi-muscle group exercise regimens and the true extent of the anti-inflammatory effects of palmar cooling may not be entirely evident. Participants in the cooling arm were also limited to the number of reps completed by their control counterparts, which may have undermined the differences in immune response between the two groups. While mass cytometry enables unprecedented resolution of the human profile of exercise and palmar cooling, the study design does not establish causality between cooling and clinical outcomes such as infection risk. Finally, the direct effect of palmar cooling on DOMS was unable to be studied as no participants met the definition for DOMS.

## CONCLUSIONS

This study provides a comprehensive analysis of the immune and metabolic responses to high intensity exercise and demonstrates that palmar cooling modulates these processes in a cell-type, pathway- and metabolite-specific manner. By targeting canonical inflammatory pathways, we showed that palmar cooling attenuated the degree of immunosuppressive responses, particularly affecting signaling pathways such as pSTAT5 and pMK2/pP38 in innate immune cells. The ability to restore immune homeostasis and reduce lactate accumulation link immune regulation with metabolic recovery. These findings highlight both the biological mechanisms and clinical potential of palmar cooling, calling for future studies assessing its value as a prophylactic strategy to preserve immune competence, as well as performance and long-term training outcomes in high-level athletes.

## Acknowledgements

We thank all volunteers for their participation in this study. Parts of Figure 1 were made using BioRender.

## Contributors

All authors have contributed to and reviewed the final manuscript. AST, VC and RL contributed equally to this paper and are joint first authors. HCH and BG contributed equally to this paper and are joint corresponding authors.

## Data availability statement

Raw data were uploaded and made publicly available on Dryad at DOI: 10.5061/dryad.n2z34tn9h. Source code is available via https://github.com/MaxSabayev/stablVMax/blob/vMax/stabl/single_omic.py

## Table and Figure Legends

**Table 1.**
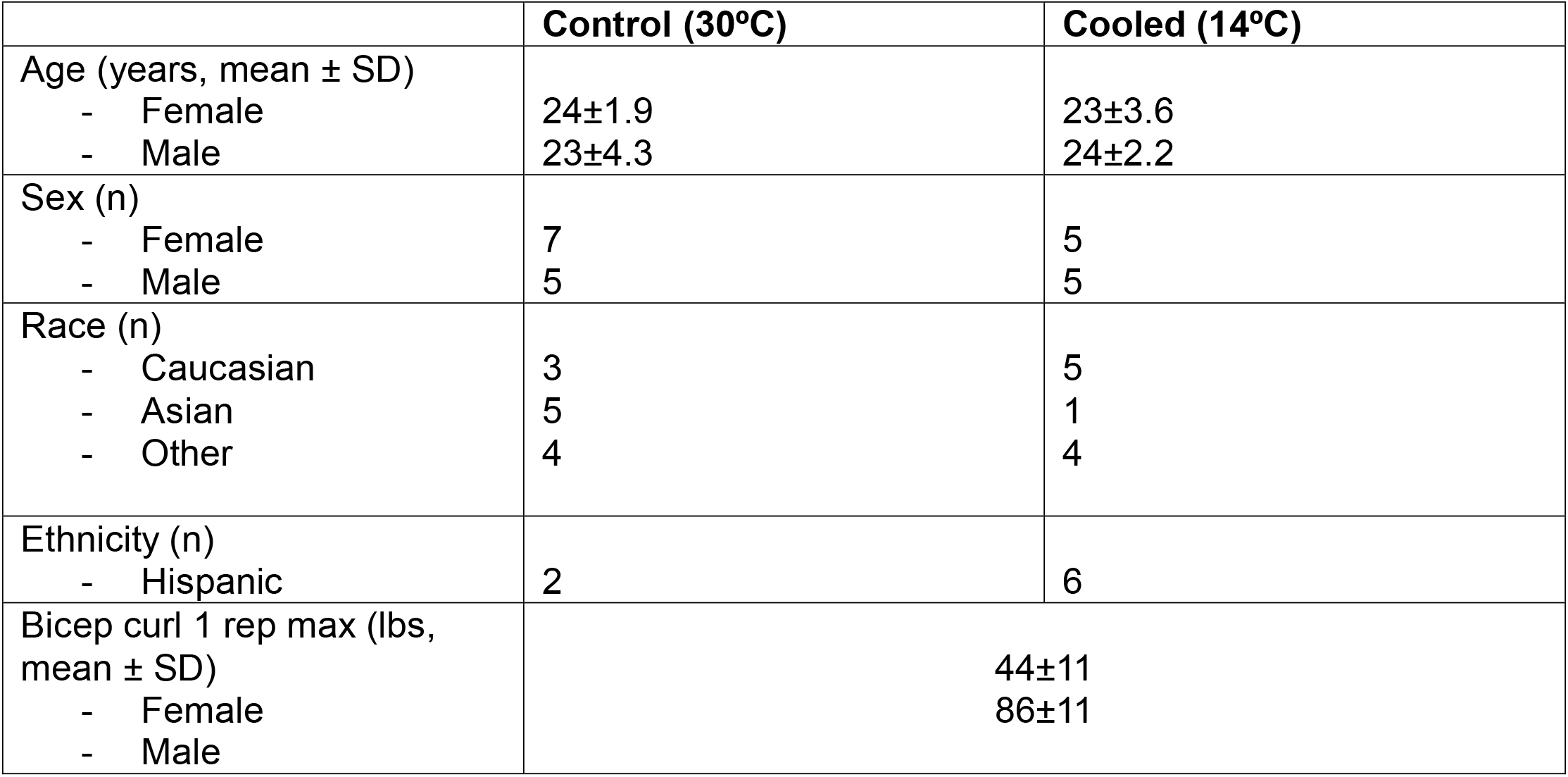
Participant demographics. There were no significant differences between control and cooled groups. Statistical significance calculated using two-sided Wilcoxon rank-sum test.

