## Supplementary Materials for "Palmar cooling mitigates exercise-induced immune suppression after high-intensity training: A randomized trial"

Supplements

Fig S1: CONSORT Map

Fig S2: Gating Strategy

Fig S3: Lactate levels

Fig S4: Pain scores

Fig S5: Frequency Trajectories in response to exercise.

Table S1: Antibody Panel

Table S2: Penalization Matrix

Table S3: Model AUC and p-values of each timepoint compared to baseline.

Table S4: All features identified in significant comparisons to baseline models.

Table S5: Model AUC and p-values of each timepoint between control and cooled arms.

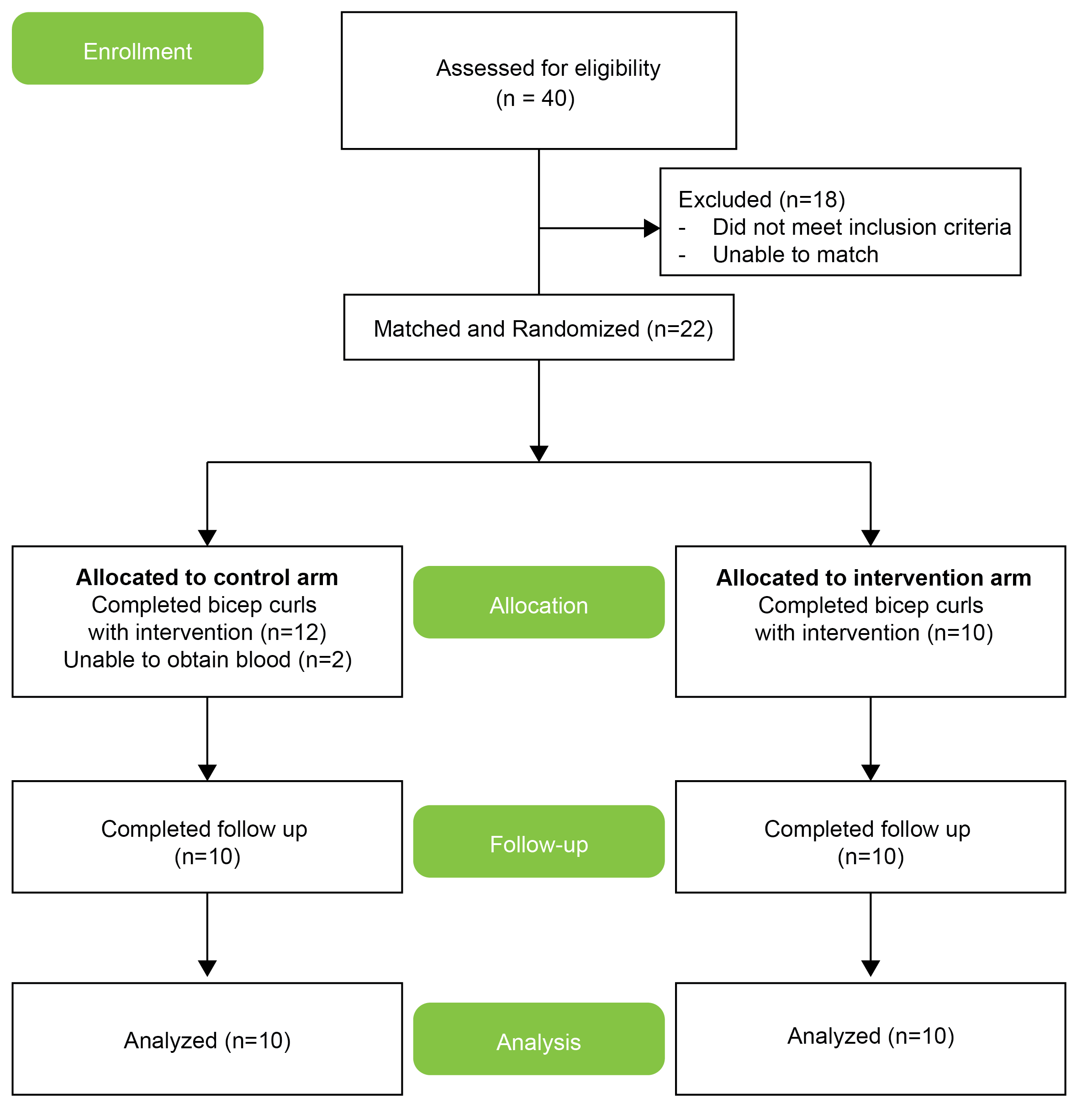

Figure S1: CONSORT Map.

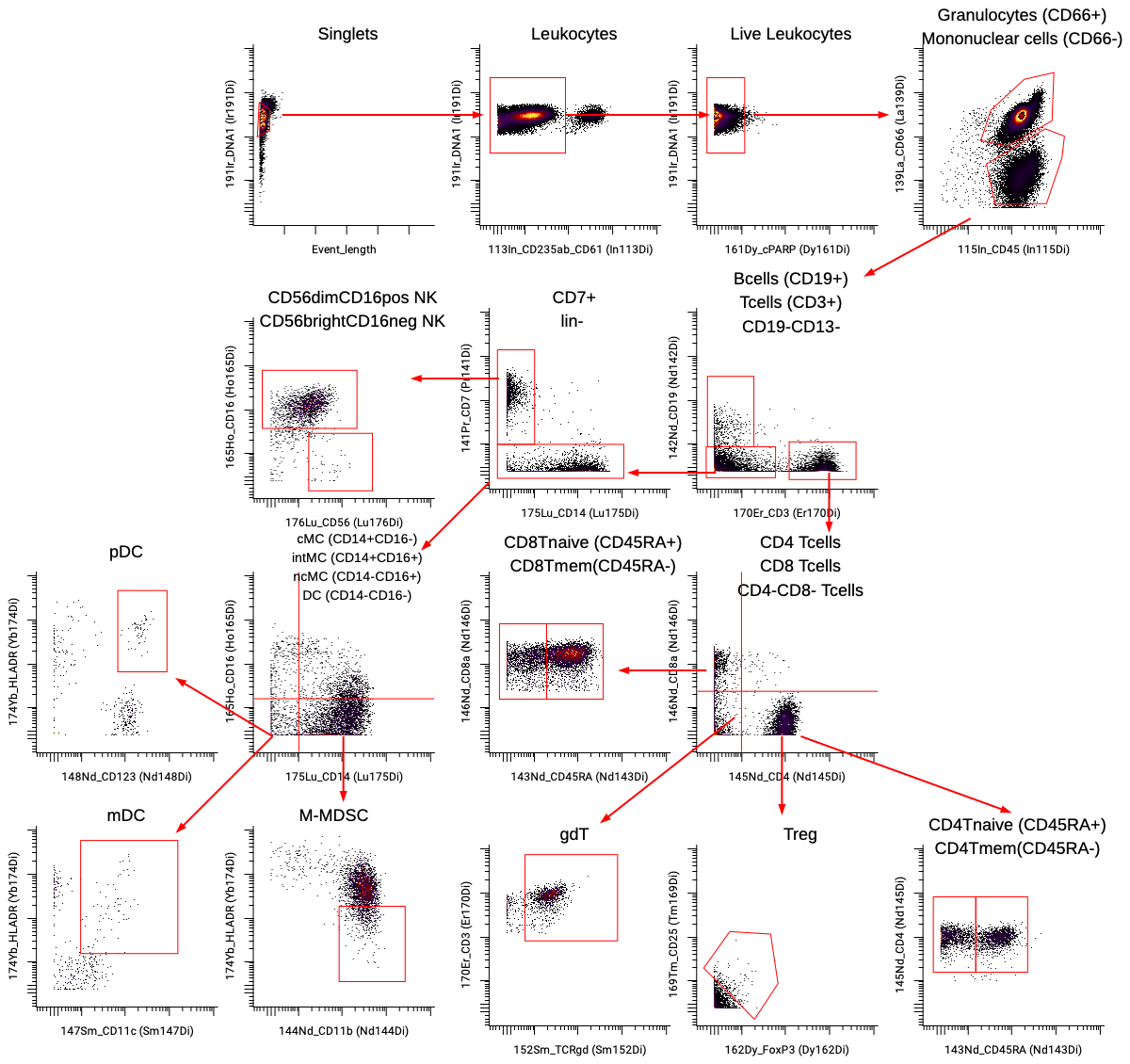

Figure S2: 2-dimensional gating strategy.

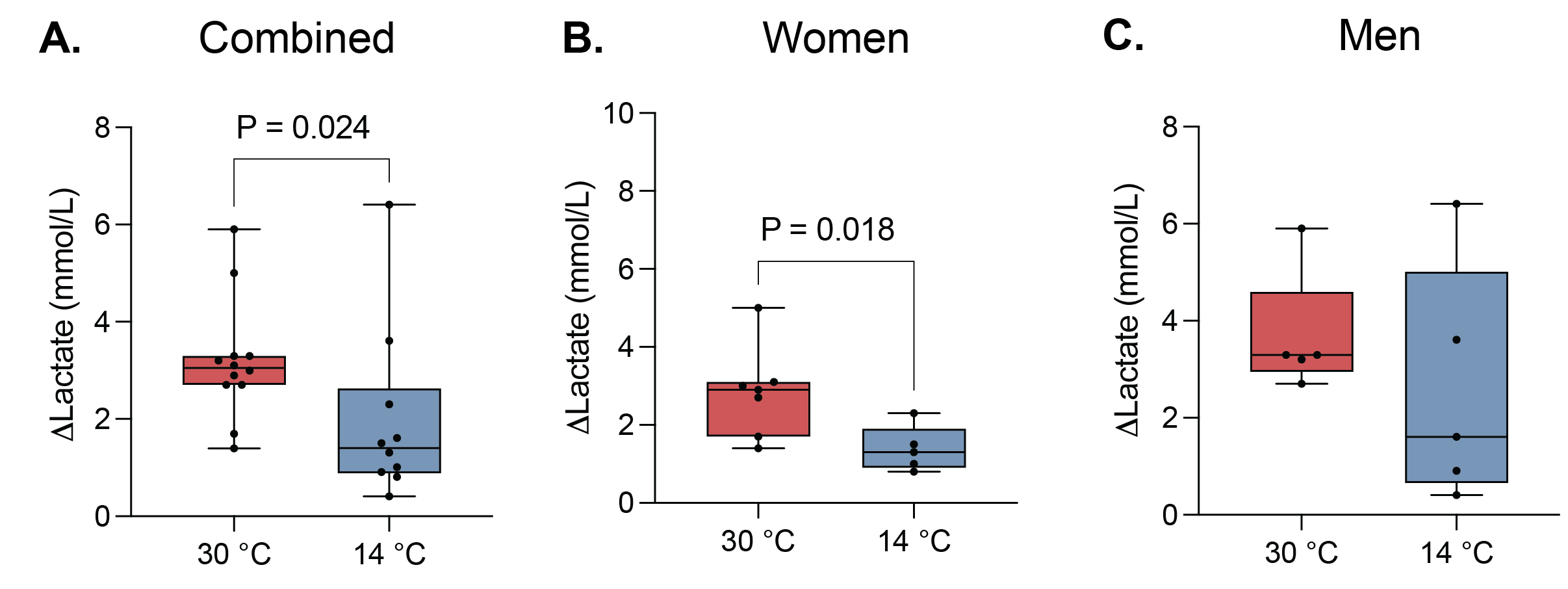

Supplemental Figure S3: . Differences in lactate levels after exercise intervention. Significant differences identified in (A) Combined cohort (p=0.024) and (B) Women (p=0.018) but not in (C) Men. Change in lactate levels are depicted in mmol/L. P-values are calculated using two-sided Wilcoxon rank-sum test. All boxplots show median values, interquartile range, whiskers of 1.5 times interquartile range.

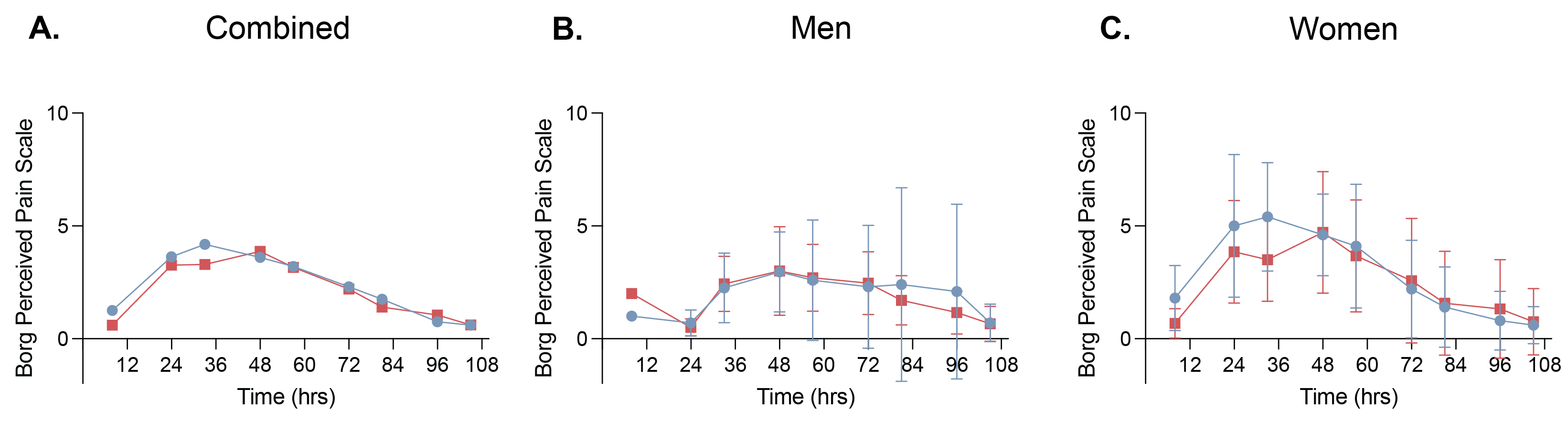
Figure S4: Pain scores reported by participants according to the Borg Perceived Pain Scale.

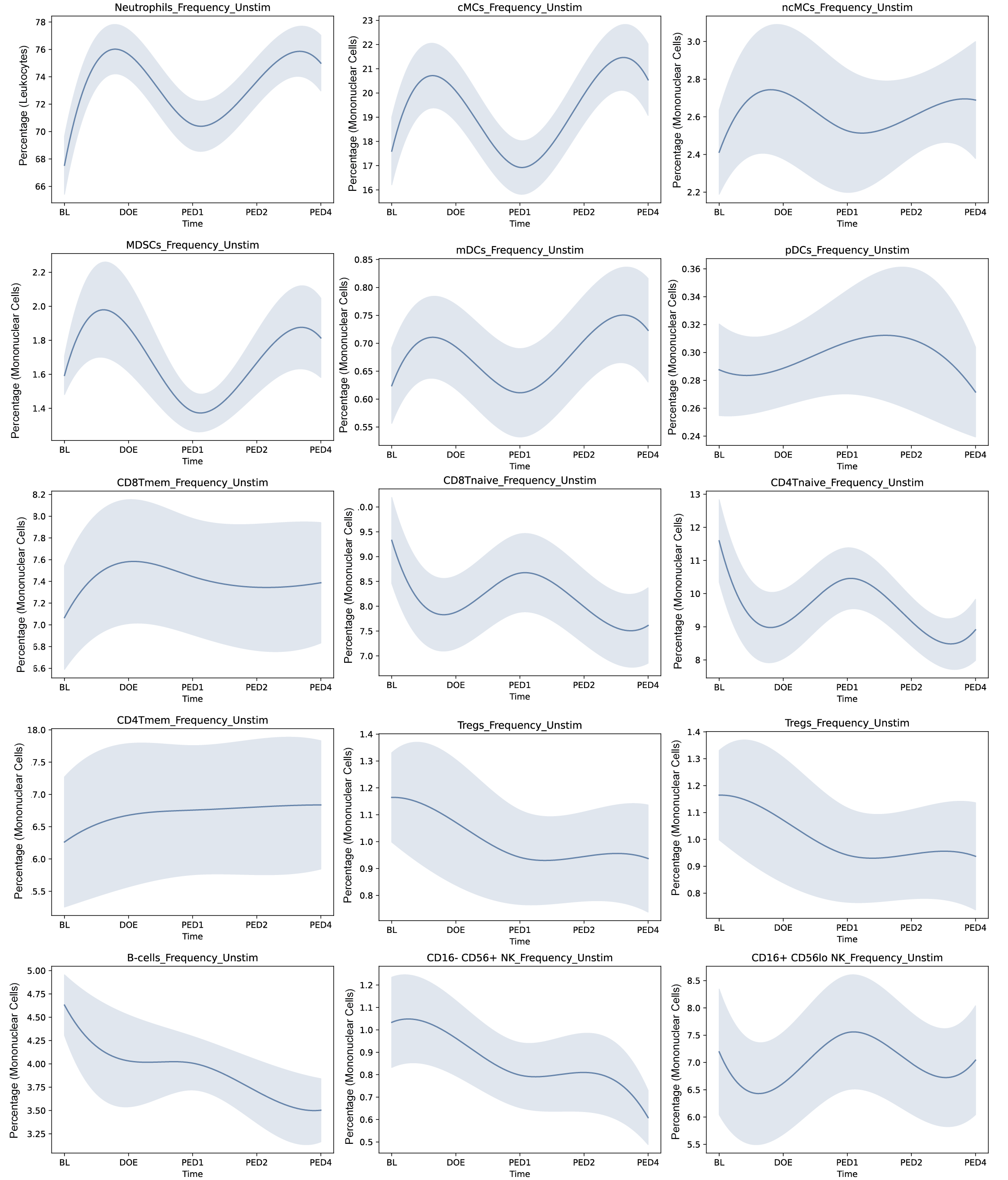
Figure S5: Frequencies of major cell types. Graph represents mean and standard error.

| Marker | Metal | Isotope | Label |
| --- | --- | --- | --- |
| Barcode | Pd | 104 | Barcode |
| Barcode | Pd | 105 | Barcode |
| Barcode | Pd | 106 | Barcode |
| Barcode | Pd | 108 | Barcode |
| Barcode | Pd | 110 | Barcode |
| CD235ab | In | 113 | Phenotypic |
| CD61 | In | 113 | Phenotypic |
| CD45 | In | 115 | Phenotypic |
| CD66 | La | 139 | Phenotypic |
| CD7 | Pr | 141 | Phenotypic |
| CD19 | Nd | 142 | Phenotypic |
| CD45RA | Nd | 143 | Phenotypic |
| CD11b | Nd | 144 | Phenotypic |
| CD4 | Nd | 145 | Phenotypic |
| CD8a | Nd | 146 | Phenotypic |
| CD11c | Sm | 147 | Phenotypic |
| CD123 | Nd | 148 | Phenotypic |
| pCREB | Sm | 149 | Functional |
| pSTAT5 | Nd | 150 | Functional |
| pp38 | Eu | 151 | Functional |
| TCRγδ | Sm | 152 | Phenotypic |
| pSTAT1 | Eu | 153 | Functional |
| pSTAT3 | Sm | 154 | Functional |
| pS6 | Gd | 155 | Functional |
| CD161 | Gd | 157 | Phenotypic |
| pMAPKAPK2 | Tb | 159 | Functional |
| Tbet | Gd | 160 | Functional |
| cPARP | Dy | 161 | Phenotypic |
| FoxP3 | Dy | 162 | Phenotypic |
| IκB | Dy | 164 | Functional |
| CD16 | Ho | 165 | Phenotypic |
| pNFκB | Er | 166 | Functional |
| pERK1/2 | Er | 167 | Functional |
| pSTAT6 | Er | 168 | Functional |
| CD25 | Tm | 169 | Phenotypic |
| CD3 | Er | 170 | Phenotypic |
| CXCR4 | Yb | 171 | Phenotypic |
| CD62L | Yb | 172 | Phenotypic |
| CCR2 | Yb | 173 | Phenotypic |
| HLA-DR | Yb | 174 | Phenotypic |
| CD14 | Yb | 175 | Phenotypic |
| CD56 | Yb | 176 | Phenotypic |
| DNA1 | Ir | 191 | DNA |
| DNA2 | Ir | 192 | DNA |
| Barcode | Pt | 194 | Barcode |
| Barcode | Pt | 195 | Barcode |
| Barcode | Pt | 196 | Barcode |
| Barcode | Pt | 197 | Barcode |

Supplemental Table 1. Staining Panel.

| **UNSTIM** | pCREB | pERK | pNFkB | IkB | pS6 | pMAPKAPK2 | pp38 | pSTAT1 | pSTAT3 | pSTAT5 | pSTAT6 |
| --- | --- | --- | --- | --- | --- | --- | --- | --- | --- | --- | --- |
| CXCR4p Neutrophils | 1 | 1 | 1 | 1 | 1 | 1 | 1 | 1 | 1 | 1 | 1 |
| Neutrophils | 1 | 1 | 1 | 1 | 1 | 1 | 1 | 1 | 1 | 1 | 1 |
| cMCs | 1 | 1 | 1 | 1 | 1 | 1 | 1 | 1 | 1 | 1 | 1 |
| MDSCs | 1 | 1 | 1 | 1 | 1 | 1 | 1 | 1 | 1 | 1 | 1 |
| mDCs | 1 | 1 | 1 | 1 | 1 | 1 | 1 | 1 | 1 | 1 | 1 |
| pDCs | 1 | 1 | 1 | 1 | 1 | 1 | 1 | 1 | 1 | 1 | 1 |
| intMCs | 1 | 1 | 1 | 1 | 1 | 1 | 1 | 1 | 1 | 1 | 1 |
| ncMCs | 1 | 1 | 1 | 1 | 1 | 1 | 1 | 1 | 1 | 1 | 1 |
| CD16- CD56+ NK | 1 | 1 | 1 | 1 | 1 | 1 | 1 | 1 | 1 | 1 | 1 |
| CD16+ CD56lo NK | 1 | 1 | 1 | 1 | 1 | 1 | 1 | 1 | 1 | 1 | 1 |
| CD4Tmem | 1 | 1 | 1 | 1 | 1 | 1 | 1 | 1 | 1 | 1 | 1 |
| CD4Tnaive | 1 | 1 | 1 | 1 | 1 | 1 | 1 | 1 | 1 | 1 | 1 |
| Tregs | 1 | 1 | 1 | 1 | 1 | 1 | 1 | 1 | 1 | 1 | 1 |
| TCRgd | 1 | 1 | 1 | 1 | 1 | 1 | 1 | 1 | 1 | 1 | 1 |
| CD8Tmem | 1 | 1 | 1 | 1 | 1 | 1 | 1 | 1 | 1 | 1 | 1 |
| CD8Tnaive | 1 | 1 | 1 | 1 | 1 | 1 | 1 | 1 | 1 | 1 | 1 |
| B-cells | 1 | 1 | 1 | 1 | 1 | 1 | 1 | 1 | 1 | 1 | 1 |
| **LPS** | pCREB | pERK | pNFkB | IkB | pS6 | pMAPKAPK2 | pp38 | pSTAT1 | pSTAT3 | pSTAT5 | pSTAT6 |
| CXCR4p Neutrophils | 1 | 1 | 1 | 1 | 1 | 1 | 1 | 0 | 0 | 0 | 0 |
| Neutrophils | 1 | 1 | 1 | 1 | 1 | 1 | 1 | 0 | 0 | 0 | 0 |
| cMCs | 1 | 1 | 1 | 1 | 1 | 1 | 1 | 0 | 0 | 0 | 0 |
| MDSCs | 1 | 1 | 1 | 1 | 1 | 1 | 1 | 0 | 0 | 0 | 0 |
| mDCs | 1 | 1 | 1 | 1 | 1 | 1 | 1 | 0 | 0 | 0 | 0 |
| pDCs | 1 | 1 | 1 | 1 | 1 | 1 | 1 | 0 | 0 | 0 | 0 |
| intMCs | 1 | 1 | 1 | 1 | 1 | 1 | 1 | 0 | 0 | 0 | 0 |
| ncMCs | 1 | 1 | 1 | 1 | 1 | 1 | 1 | 0 | 0 | 0 | 0 |
| CD16- CD56+ NK | 1 | 1 | 1 | 1 | 1 | 1 | 1 | 0 | 0 | 0 | 0 |
| CD16+ CD56lo NK | 1 | 1 | 1 | 1 | 1 | 1 | 1 | 0 | 0 | 0 | 0 |
| CD4Tmem | 0 | 0 | 0 | 0 | 0 | 0 | 0 | 0 | 0 | 0 | 0 |
| CD4Tnaive | 0 | 0 | 0 | 0 | 0 | 0 | 0 | 0 | 0 | 0 | 0 |
| Tregs | 1 | 1 | 1 | 1 | 1 | 1 | 1 | 0 | 0 | 0 | 0 |
| TCRgd | 0 | 0 | 0 | 0 | 0 | 0 | 0 | 0 | 0 | 0 | 0 |
| CD8Tmem | 0 | 0 | 0 | 0 | 0 | 0 | 0 | 0 | 0 | 0 | 0 |
| CD8Tnaive | 0 | 0 | 0 | 0 | 0 | 0 | 0 | 0 | 0 | 0 | 0 |
| B-cells | 0 | 0 | 0 | 0 | 0 | 0 | 0 | 0 | 0 | 0 | 0 |
| **IFNa** | pCREB | pERK | pNFkB | IkB | pS6 | pMAPKAPK2 | pp38 | pSTAT1 | pSTAT3 | pSTAT5 | pSTAT6 |
| CXCR4p Neutrophils | 0 | 0 | 0 | 0 | 0 | 0 | 1 | 1 | 1 | 1 | 1 |
| Neutrophils | 0 | 0 | 0 | 0 | 0 | 0 | 1 | 1 | 1 | 1 | 1 |
| cMCs | 0 | 0 | 0 | 0 | 0 | 0 | 1 | 1 | 1 | 1 | 1 |
| MDSCs | 0 | 0 | 0 | 0 | 0 | 0 | 1 | 1 | 1 | 1 | 1 |
| mDCs | 0 | 0 | 0 | 0 | 0 | 0 | 1 | 1 | 1 | 1 | 1 |
| pDCs | 0 | 0 | 0 | 0 | 0 | 0 | 1 | 1 | 1 | 1 | 1 |
| intMCs | 0 | 0 | 0 | 0 | 0 | 0 | 1 | 1 | 1 | 1 | 1 |
| ncMCs | 0 | 0 | 0 | 0 | 0 | 0 | 1 | 1 | 1 | 1 | 1 |
| CD16- CD56+ NK | 0 | 0 | 0 | 0 | 0 | 0 | 1 | 1 | 1 | 1 | 1 |
| CD16+ CD56lo NK | 0 | 0 | 0 | 0 | 0 | 0 | 1 | 1 | 1 | 1 | 1 |
| CD4Tmem | 0 | 0 | 0 | 0 | 0 | 0 | 1 | 1 | 1 | 1 | 1 |
| CD4Tnaive | 0 | 0 | 0 | 0 | 0 | 0 | 1 | 1 | 1 | 1 | 1 |
| Tregs | 0 | 0 | 0 | 0 | 0 | 0 | 1 | 1 | 1 | 1 | 1 |
| TCRgd | 0 | 0 | 0 | 0 | 0 | 0 | 1 | 1 | 1 | 1 | 1 |
| CD8Tmem | 0 | 0 | 0 | 0 | 0 | 0 | 1 | 1 | 1 | 1 | 1 |
| CD8Tnaive | 0 | 0 | 0 | 0 | 0 | 0 | 1 | 1 | 1 | 1 | 1 |
| B-cells | 0 | 0 | 0 | 0 | 0 | 0 | 1 | 1 | 1 | 1 | 1 |

Table S2. Penalization Matrix. 1 indicates weighted features, 0 indicates unweighted features.

|  | Day of Exercise | Post-exercise Day 1 | Post-exercise Day 2 | Post-exercise Day 4 |
| --- | --- | --- | --- | --- |
| AUC | 0.67 | 0.80 | 0.69 | 0.69 |
| p-val | 0.064 | 0.0015 | 0.041 | 0.040 |

Table S3. Comparison to baseline.

| **PED1** | **PED2** | | **PED4** |
| --- | --- | --- | --- |
| B-cells_Frequency_Unstim | B-cells_pp38_IFNa | mDCs_pCREB_Unstim | B-cells_pp38_IFNa |
| CD16- CD56+ NK_Frequency_Unstim | CD16- CD56+ NK_IkB_LPS | mDCs_pp38_IFNa | intMCs_pERK_LPS |
| CXCR4p Neutrophils_pCREB_LPS | CD16- CD56+ NK_IkB_Unstim | mDCs_pp38_LPS | ncMCs_pCREB_LPS |
| intMCs_pERK_LPS | CD16- CD56+ NK_pERK_LPS | mDCs_pS6_LPS | ncMCs_pp38_IFNa |
| MDSCs_Frequency_Unstim | CD16- CD56+ NK_pMAPKAPK2_LPS | mDCs_pSTAT3_IFNa |  |
| MDSCs_pMAPKAPK2_LPS | CD16- CD56+ NK_pp38_IFNa | MDSCs_IkB_LPS |  |
| ncMCs_Frequency_Unstim | CD16- CD56+ NK_pSTAT5_IFNa | MDSCs_pS6_Unstim |  |
| ncMCs_pMAPKAPK2_LPS | CD16- CD56+ NK_pSTAT5_Unstim | ncMCs_IkB_LPS |  |
| ncMCs_pp38_LPS | CD16+ CD56lo NK_Frequency_Unstim | ncMCs_pCREB_LPS |  |
| Neutrophils_Frequency_Unstim | CD16+ CD56lo NK_IkB_LPS | ncMCs_pp38_IFNa |  |
| Neutrophils_pp38_LPS | CD16+ CD56lo NK_pCREB_LPS | ncMCs_pp38_LPS |  |
| pDCs_pS6_LPS | CD16+ CD56lo NK_pERK_LPS | Neutrophils_Frequency_Unstim |  |
| Tregs_Frequency_Unstim | CD16+ CD56lo NK_pSTAT1_IFNa | Neutrophils_IkB_LPS |  |
|  | CD16+ CD56lo NK_pSTAT3_IFNa | Neutrophils_pS6_LPS |  |
|  | CD16+ CD56lo NK_pSTAT5_IFNa | pDCs_IkB_LPS |  |
|  | CD4Tmem_pp38_LPS | pDCs_IkB_Unstim |  |
|  | CD4Tnaive_pp38_IFNa | pDCs_pMAPKAPK2_LPS |  |
|  | CD4Tnaive_pSTAT3_IFNa | pDCs_pSTAT3_Unstim |  |
|  | CD4Tnaive_pSTAT5_IFNa | Tregs_pERK_LPS |  |
|  | CD8Tmem_pSTAT5_Unstim | Tregs_pMAPKAPK2_LPS |  |
|  | CXCR4p Neutrophils_Frequency_Unstim | Tregs_pS6_Unstim |  |
|  | intMCs_pS6_Unstim | Tregs_pSTAT5_Unstim |  |
|  | mDCs_IkB_LPS |  |  |

Table S4. Features selected by each significant comparison to baseline model depicted as cell population_marker_stimulation. Ranked by alphabetical order.

|  | **Baseline** | **Day of Exercise** | **Post-exercise Day 1** | **Post-exercise Day 2** | **Post-exercise Day 4** |
| --- | --- | --- | --- | --- | --- |
| AUC | 0.63 | 0.58 | 0.59 | 0.79 | 0.71 |
| p-val | 0.34 | 0.57 | 0.52 | 0.031 | 0.12 |

Table S5. Comparison between Cooled and Control arms.
